# Active safety surveillance of COVID-19 mRNA vaccines in children aged 5-15 years in Australia

**DOI:** 10.1101/2022.07.19.22277827

**Authors:** Nicholas Wood, Laura K Lopez, Catherine Glover, Alan Leeb, Patrick Cashman, Lucy Deng, Kristine Macartney

## Abstract

AusVaxSafety (Australia’s active safety surveillance system) used SMS/email delivered surveys to actively solicit the short-term (within first 3 days after vaccination) adverse event profile of mRNA COVID-19 vaccines in children (aged 5–15 years) by age, dose, brand and pre-existing comorbidity.

392,268 survey responses for children aged 5–15 who received a COVID-19 vaccine between July 2021–May 2022 (211,994 following BNT162b2 10mcg in children aged 5–11 years, 173,329 following BNT162b2 30mcg and 6,945 following mRNA-1273 100mcg in adolescents aged 12–15 years) were analysed.

Adverse event rates were higher following dose 2 and 3 compared to dose 1 after all vaccines and highest following dose 2 of mRNA-1273 in 12–15 years. Fever was low in the youngest children (5 years old, any dose; 1,090/26,181 (4%)). Medical review rates remained low (0.3% overall) and impact on daily activities was also low (7% overall). No self-reported cases of myocarditis or pericarditis were identified.

Ongoing active safety surveillance of lower dose mRNA vaccines in children under 5 years old is required to better understand safety as the vaccines roll out into this population age-group.

## INTRODUCTION

Australia commenced its COVID-19 vaccine program for children on 10 January 2022 with children aged 5–11 years recommended to receive two doses of Comirnaty (Pfizer–BioNTech BNT162b2, 10 micrograms) separated by an 8 week interval. From 24 February 2022, children aged 6–11 years could receive two doses of Spikevax (Moderna mRNA-1273, 50 micrograms), 8 weeks apart.

Until recently Australia has been one of the few countries globally to administer mRNA-1273 to children under 12 years old. From 18 June, the US has recommended either BNT162b2 (3 micrograms) or mRNA-1273 (25 micrograms) in children aged 6 months-5 years. (1)

There is limited safety data from clinical trials in children aged 5-11 years for BNT162b2 (2) and mRNA-1273. (3) US V-safe, (4) a voluntary smartphone-based active COVID-19 vaccine safety surveillance system, has reported on the short-term safety of the BNT162b2 doses (3 weeks apart) in children aged 5-11 years, but not mRNA-1273. (5)

Australia’s active safety surveillance system AusVaxSafety monitors the safety of COVID-19 vaccines. We report on the short-term adverse event profile of mRNA COVID-19 vaccines in >300,000 children (aged 5–15 years) by age, dose, brand and pre-existing comorbidity.

## METHODS

Children aged 5–15 years who received a COVID-19 vaccine at vaccination sites (state-run vaccination hubs, pharmacy, primary healthcare practices) were included in our analysis, with active prospective survey-based methods as previously described. (6)

Three days after vaccination, the child’s parent/guardian was sent an SMS or email with a link to an online survey with defined response options about adverse events following immunisation (AEFI), any medical care or advice sought and impact on daily activities. Solicited local adverse events and systemic adverse events included myalgia, arthralgia, headache, fever, chills, fatigue and gastrointestinal symptoms. Vaccination details (vaccine brand, batch, dose, date) and demographic details (age, sex, Indigenous status, underlying medical conditions) were obtained. (6)

We examined the proportion of respondents reporting any AEFI 0–3 days post-vaccination, reporting medical review for AEFI and impact on daily activities. All analyses were conducted in R version 4.1.0. (7) The study had ethics approval at the Sydney Children’s Hospitals Network (HREC/16/SCHN/19).

## RESULTS

We received 392,268 survey responses for children aged 5–15 who received a COVID-19 vaccine between July 2021–May 2022; 211,994 following BNT162b2 10mcg in children aged 5–11 years, 173,329 following BNT162b2 30mcg and 6,945 following mRNA-1273 100mcg in 12–15 years (Table 1). Only 60 reports for mRNA-1273 in children aged 6–11 years were received precluding meaningful analysis and therefore were excluded. AEFI rates were similar across sex and Indigenous status but higher for children with parent-reported chronic medical conditions. Medical review rates remained low (0.3% overall) and impact on daily activities was also low (7% overall).

**Table 1:** Adverse event rates reported by age, sex, underlying medical conditions and vaccine brand and dose.

|  | Comirnaty<br>(Pfizer, 10 micrograms)<br>5–11 years |  |  | Comirnaty<br>(Pfizer, 30 micrograms)<br>12–15 years |  |  | Spikevax<br>(Moderna, 100 micrograms)<br>12–15 years |  |  |
| --- | --- | --- | --- | --- | --- | --- | --- | --- | --- |
|  | Dose 1<sup>1</sup> | Dose 2<sup>1</sup> | Dose 3<sup>1</sup> | Dose 1<sup>1</sup> | Dose 2<sup>1</sup> | Dose 3<sup>1</sup> | Dose 1<sup>1</sup> | Dose 2<sup>1</sup> | Dose 3<sup>1</sup> |
| **Adverse event rate (all participants)** | 33,597/132,313 (25%) | 22,115/79,542 (28%) | 17/59 (29%) | 30,217/94,527 (32%) | 38,608/78,158 (49%) | 320/644 (50%) | 1,213/3,569 (34%) | 2,116/3,319 (64%) | 38/57 (67%) |
| **Adverse event rate by participant characteristics** |  |  |  |  |  |  |  |  |  |
| **Sex** |  |  |  |  |  |  |  |  |  |
| Female | 17,539/63,572 (28%) | 11,350/38,290 (30%) | 10/27 (37%) | 15,427/47,218 (33%) | 19,371/38,747 (50%) | 145/288 (50%) | 489/1,421 (34%) | 764/1,213 (63%) | 20/24 (83%) |
| Male | 15,613/66,801 (23%) | 10,397/39,828 (26%) | 5/27 (19%) | 14,627/46,758 (31%) | 18,995/38,913 (49%) | 163/313 (52%) | 426/1,329 (32%) | 765/1,183 (65%) | 11/17 (65%) |
| Another Term | 25/88 (28%) | 15/36 (42%) | 0/0 (NA%) | 74/234 (32%) | 82/167 (49%) | 1/2 (50%) | 0/1 (0%) | 2/2 (100%) | 0/0 (NA%) |
| Unknown | 420/1,852 (23%) | 353/1,388 (25%) | 2/5 (40%) | 89/317 (28%) | 160/331 (48%) | 11/41 (27%) | 298/818 (36%) | 585/921 (64%) | 7/16 (44%) |
| **Indigenous status** |  |  |  |  |  |  |  |  |  |
| Aboriginal and Torres Strait Islander | 787/3,338 (24%) | 500/1,956 (26%) | 0/1 (0%) | 1,021/3,349 (30%) | 1,100/2,540 (43%) | 14/27 (52%) | 41/112 (37%) | 47/91 (52%) | 1/2 (50%) |
| Not indigenous | 31,850/124,633 (26%) | 21,263/76,101 (28%) | 17/57 (30%) | 28,614/89,125 (32%) | 36,833/74,086 (50%) | 301/598 (50%) | 1,148/3,381 (34%) | 2,037/3,167 (64%) | 36/54 (67%) |
| **Children with history of anaphylaxis** | 1,000/3,222 (31%) | 611/2,002 (31%) | 2/7 (29%) | 1,075/2,952 (36%) | 1,413/2,462 (57%) | 14/35 (40%) | 25/70 (36%) | 44/71 (62%) | 1/1 (100%) |
| **Chronic medical condition** |  |  |  |  |  |  |  |  |  |
| Yes | 1,337/3,674 (36%) | 800/2,065 (39%) | 11/33 (33%) | 1,665/3,708 (45%) | 1,716/2,745 (63%) | 106/202 (52%) | 39/81 (48%) | 59/77 (77%) | 5/8 (62%) |
| No | 32,260/128,639 (25%) | 21,315/77,477 (28%) | 6/26 (23%) | 28,552/90,819 (31%) | 36,892/75,413 (49%) | 214/442 (48%) | 1,174/3,488 (34%) | 2,057/3,242 (63%) | 33/49 (67%) |
| **Specific chronic medical condition<sup>2</sup>** |  |  |  |  |  |  |  |  |  |
| Chronic cardiorespiratory, renal or hepatic disease<sup>3</sup> | 209/662 (32%) | 122/360 (34%) | 1/6 (17%) | 363/874 (42%) | 351/611 (57%) | 18/29 (62%) | 4/13 (31%) | 5/9 (56%) | 2/3 (67%) |
| Diabetes | 76/229 (33%) | 49/125 (39%) | 0/2 (0%) | 150/388 (39%) | 157/292 (54%) | 6/11 (55%) | 1/8 (12%) | 2/5 (40%) | 1/2 (50%) |
| Haematological/Oncological condition | 36/136 (26%) | 27/97 (28%) | 6/16 (38%) | 28/89 (31%) | 40/87 (46%) | 19/39 (49%) | 0/0 (NA%) | 0/0 (NA%) | 0/1 (0%) |
| Neurological condition | 81/233 (35%) | 49/141 (35%) | 1/3 (33%) | 88/202 (44%) | 79/148 (53%) | 6/13 (46%) | 0/0 (NA%) | 1/2 (50%) | 2/2 (100%) |
| Immunological condition | 61/163 (37%) | 43/125 (34%) | 2/9 (22%) | 123/268 (46%) | 134/212 (63%) | 31/58 (53%) | 5/6 (83%) | 3/3 (100%) | 2/2 (100%) |
| Obesity | 11/32 (34%) | 5/19 (26%) | 0/0 (NA%) | 27/53 (51%) | 23/35 (66%) | 2/4 (50%) | 1/1 (100%) | 2/3 (67%) | 0/0 (NA%) |
| Other<sup>4</sup> | 1,016/2,684 (38%) | 602/1,457 (41%) | 3/6 (50%) | 1,149/2,465 (47%) | 1,182/1,798 (66%) | 45/88 (51%) | 31/62 (50%) | 48/61 (79%) | 0/1 (0%) |
| **Medical Review** | 701/132,313 (0.5%) | 570/79,542 (0.7%) | 0/59 (0%) | 572/94,527 (0.6%) | 1,111/78,158 (1%) | 13/644 (2%) | 35/3,569 (1.3%) | 103/3,319 (3.1%) | 1/57 (1.7%) |
| **Highest level of medical review** |  |  |  |  |  |  |  |  |  |
| Phone | 175/701 (25%) | 149/570 (26%) | 0/0 (NA%) | 113/572 (20%) | 237/1,111 (21%) | 4/13 (31%) | 6/35 (17%) | 27/103 (26%) | 0/1 (0%) |
| Primary healthcare<sup>5</sup> | 328/701 (47%) | 273/570 (48%) | 0/0 (NA%) | 265/572 (46%) | 483/1,111 (43%) | 3/13 (23%) | 6/35 (17%) | 41/103 (40%) | 1/1 (100%) |
| ED<sup>6</sup> | 166/701 (24%) | 115/570 (20%) | 0/0 (NA%) | 159/572 (28%) | 355/1,111 (32%) | 6/13 (46%) | 15/35 (43%) | 23/103 (22%) | 0/1 (0%) |
| Unknown | 32/701 (4.6%) | 33/570 (5.8%) | 0/0 (NA%) | 35/572 (6.1%) | 36/1,111 (3.2%) | 0/13 (0%) | 8/35 (23%) | 12/103 (12%) | 0/1 (0%) |
| **Impact on daily activities<sup>7</sup>** | 3,977/132,313 (3%) | 5,866/79,542 (7%) | 4/59 (7%) | 5,761/94,527 (6%) | 17,512/78,158 (22%) | 126/644 (19%) | 312/3,569 (8.7%) | 1,274/3,319 (38%) | 17/57 (29%) |
| **Number of days impacted** |  |  |  |  |  |  |  |  |  |
| < 1 day | 598/3,977 (15%) | 679/5,866 (12%) | 0/4 (0%) | 918/5,761 (16%) | 2,233/17,512 (13%) | 11/126 (8.7%) | 26/312 (8.3%) | 83/1,274 (3.9%) | 1/17 (2.6%) |
| 1 day | 1,984/3,977 (50%) | 3,136/5,866 (54%) | 3/4 (75%) | 2,925/5,761 (51%) | 9,719/17,512 (55%) | 48/126 (38%) | 182/312 (58%) | 694/1,274 (33%) | 12/17 (32%) |
| 2 days | 1,002/3,977 (25%) | 1,418/5,866 (24%) | 1/4 (25%) | 1,476/5,761 (26%) | 4,448/17,512 (25%) | 45/126 (36%) | 71/312 (23%) | 377/1,274 (18%) | 4/17 (11%) |
| >= 3 days | 391/3,977 (9.8%) | 629/5,866 (11%) | 0/4 (0%) | 438/5,761 (7.6%) | 1,107/17,512 (6.3%) | 22/126 (17%) | 33/312 (11%) | 119/1,274 (5.6%) | 0/17 (0%) |

<sup>1</sup>n/N (%); <sup>2</sup>Sum of denominators of medical condition groups exceeds the total denominator of chronic medical condition as respondents may select more than one condition; <sup>3</sup>Includes heart disease, poorly controlled hypertension, chronic lung disease, chronic kidney disease, chronic liver disease; <sup>4</sup>Other includes autoimmune conditions, allergies and sleep disorders, among others; <sup>5</sup>Primary healthcare includes general practitioner or Aboriginal healthcare worker; <sup>6</sup>Visit to a hospital emergency department; <sup>7</sup>Impact defined as vaccination event causing respondent to miss work, study, normal duties or routine.

The proportion of children with a reported AEFI (any, local or systemic) trended upwards with age (Figure 1). AEFI rates were higher following dose 2 and 3 compared to dose 1 (Supplementary Table 1 and Supplementary Figure 1), though the difference between dose 1 and 2 was much lower in 5–11 years than in 12–15 years. AEFI rates were highest following dose 2 of mRNA-1273 in 12–15 years and impact on routine activities was also more common after dose 2 or 3 of either vaccine in this age group (Table 1). No self-reported cases of myocarditis or pericarditis were identified.

**Figure 1.**
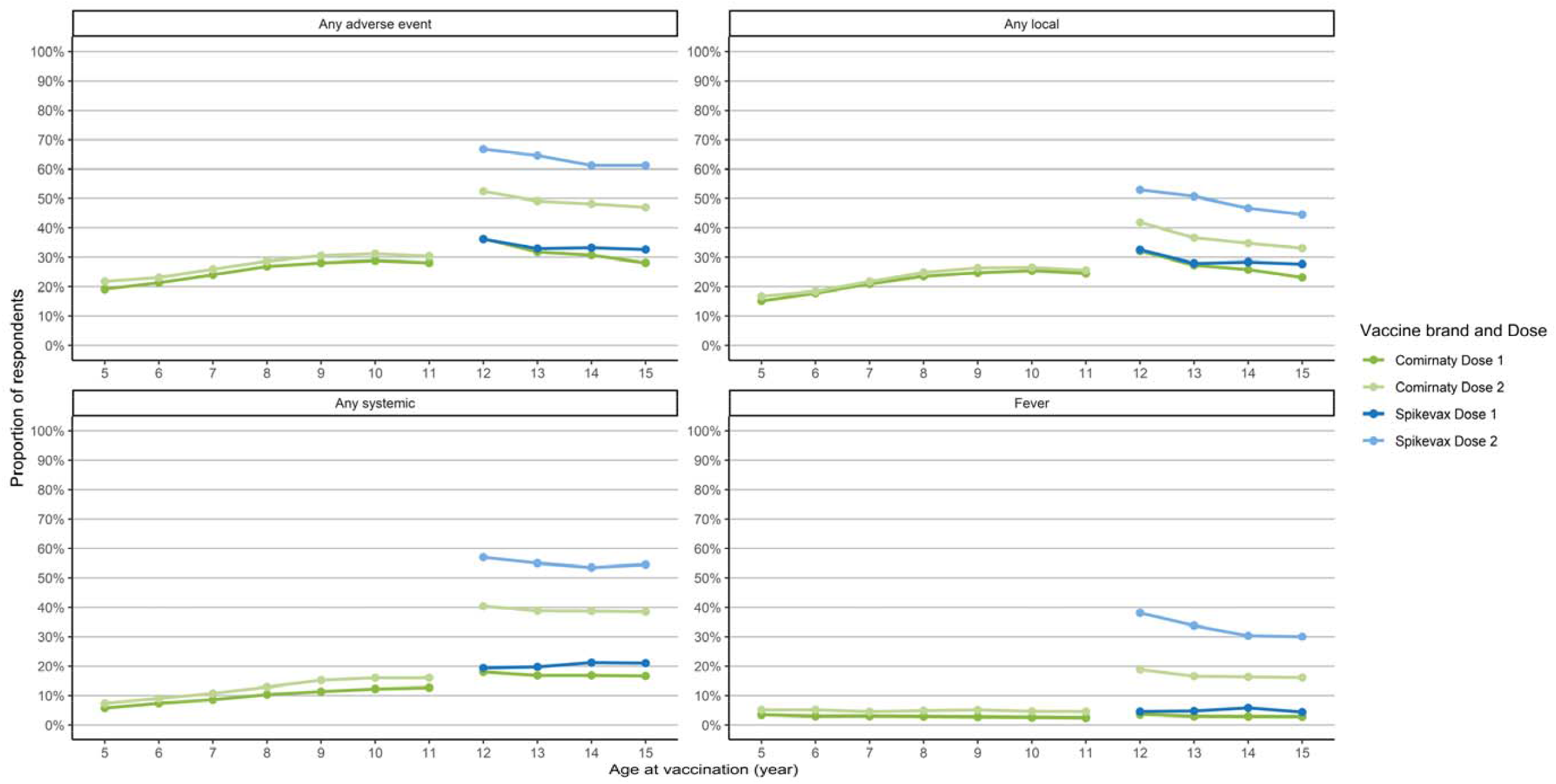
Adverse event rates by age year, vaccine brand*, and dose. *Dose 3 data for Comirnaty 5-11, Comirnaty 12-15 and Spikevax 12-15 excluded from figure due to small number of responses.

## DISCUSSION

Active safety surveillance data from Australia found lower AEFI reporting rates in the days following vaccination of both mRNA vaccines in children aged 5–15 years compared to known short-term safety data. (2,3,5) AEFI reporting rates were lower in younger children (aged 5–11 years) compared to older children potentially related to the use of reduced dose mRNA vaccines. Adverse event rates were slightly higher in children with chronic medical conditions, which may reflect closer observation by their parents. Importantly fever, which is a concern in children under 5 years old due to the potential for febrile seizures, was low in the youngest children (5 years, any dose; 1,090/26,181 (4%)) and similar to that seen following annual influenza vaccination. (8) While data reflects survey respondents only, those with adverse events are possibly more likely to complete the survey. These data should therefore provide confidence to parents in the safety of COVID-19 vaccines; active safety surveillance of lower dose mRNA vaccines in children under 5 years old is needed to better understand safety in this population.

## Data Availability

All data produced in the present study are available upon reasonable request to the authors

## Abbreviations

(AEFI): Adverse event following immunisation

## ACKNOWLEDGEMENTS

AusVaxSafety surveillance is funded under a contract with the Australian Department of Health. We acknowledge the participants and staff at the surveillance sites, state and territory health departments, and the contribution of the surveillance tools SmartVax, Vaxtracker, and Microsoft COVID Vaccine Management System.

## TABLES

Supplementary Table 1: Solicited adverse event rates by age, dose and vaccine brand

## FIGURES

Supplementary Figure 1. Solicited adverse event rates by vaccine formulation and dose number

