## Supplementary figures and images for "Active safety surveillance of COVID-19 mRNA vaccines in children aged 5-15 years in Australia"

### Supplemental Figure 1

**Supplementary Figure 1. Solicited adverse event rates by vaccine formulation and dose number**

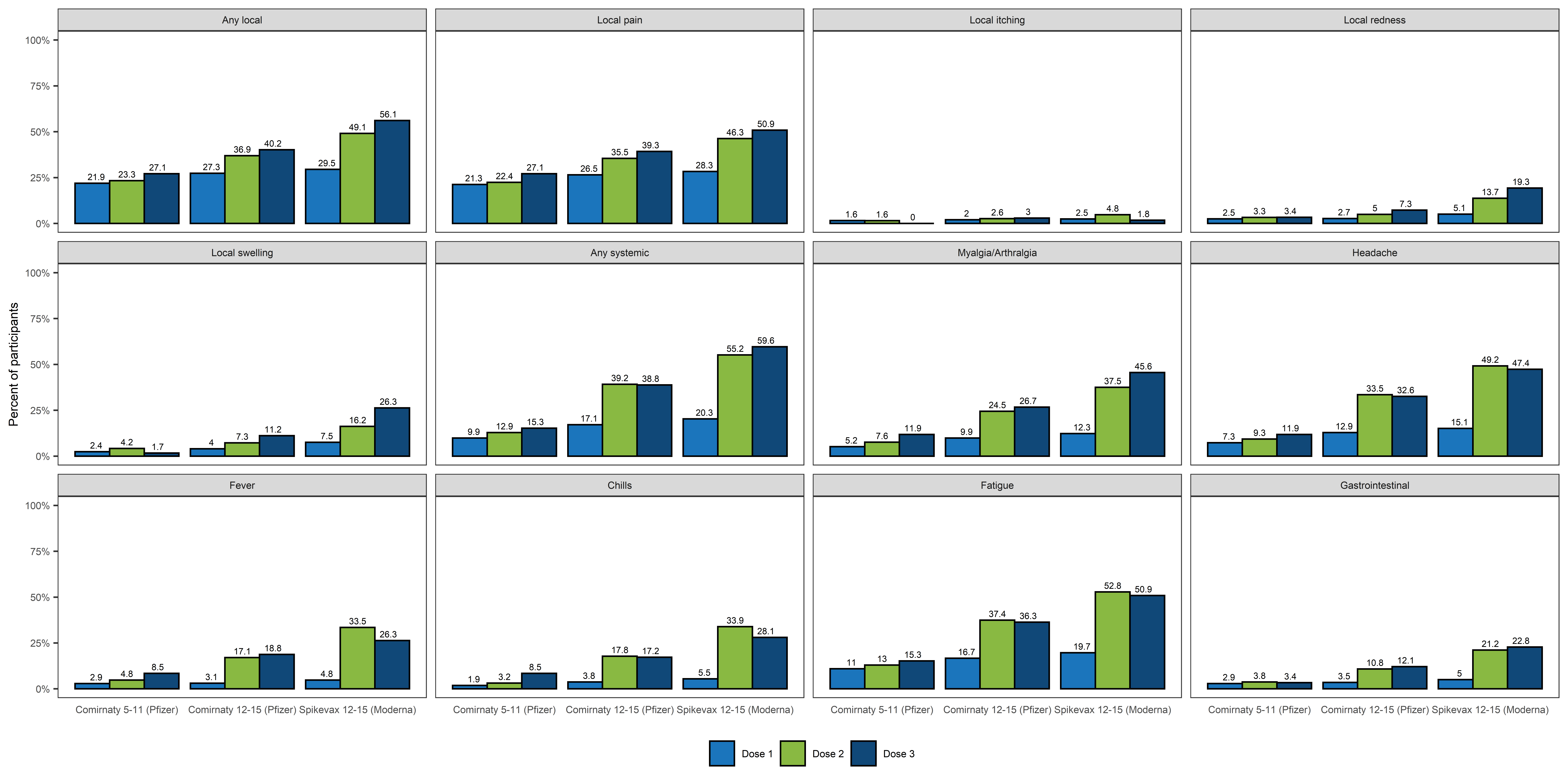
