## Supplemental Table 1 for "Active safety surveillance of COVID-19 mRNA vaccines in children aged 5-15 years in Australia"

**Supplementary Table 1: Solicited adverse event rates by age, dose and vaccine brand**

|  | **Comirnaty (Pfizer, 10 micrograms)**  **5–11 years** | | | **Comirnaty (Pfizer, 30 micrograms)**  **12–15 years** | | | **Spikevax (Moderna, 100 micrograms)**  **12–15 years** | | |
| --- | --- | --- | --- | --- | --- | --- | --- | --- | --- |
| **Adverse Event** | **Dose 1**  **(N = 132313)^1^** | **Dose 2**  **(N = 79542)^1^** | **Dose 3**  **(N = 59)^1^** | **Dose 1**  **(N = 94527)^1^** | **Dose 2**  **(N = 78158)^1^** | **Dose 3**  **(N = 644)^1^** | **Dose 1**  **(N = 3569)^1^** | **Dose 2**  **(N = 3319)^1^** | **Dose 3**  **(N = 57)^1^** |
| **Local** | 28,997 (22%) | 18,560 (23%) | 16 (27%) | 25,852 (27%) | 28,879 (37%) | 259 (40%) | 1,052 (29%) | 1,630 (49%) | 32 (56%) |
| Local pain | 28,132 (21%) | 17,817 (22%) | 16 (27%) | 25,063 (27%) | 27,715 (35%) | 253 (39%) | 1,011 (28%) | 1,537 (46%) | 29 (51%) |
| Local itching | 2,092 (1.6%) | 1,236 (1.6%) | 0 (0%) | 1,913 (2.0%) | 2,055 (2.6%) | 19 (3.0%) | 88 (2.5%) | 158 (4.8%) | 1 (1.8%) |
| Local redness | 3,361 (2.5%) | 2,654 (3.3%) | 2 (3.4%) | 2,557 (2.7%) | 3,895 (5.0%) | 47 (7.3%) | 181 (5.1%) | 456 (14%) | 11 (19%) |
| Local swelling | 3,236 (2.4%) | 3,341 (4.2%) | 1 (1.7%) | 3,821 (4.0%) | 5,672 (7.3%) | 72 (11%) | 269 (7.5%) | 539 (16%) | 15 (26%) |
| **Systemic** | 13,066 (9.9%) | 10,234 (13%) | 9 (15%) | 16,208 (17%) | 30,644 (39%) | 250 (39%) | 725 (20%) | 1,832 (55%) | 34 (60%) |
| Myalgia/Arthralgia | 6,859 (5.2%) | 6,039 (7.6%) | 7 (12%) | 9,367 (9.9%) | 19,144 (24%) | 172 (27%) | 440 (12%) | 1,246 (38%) | 26 (46%) |
| Headache | 9,648 (7.3%) | 7,407 (9.3%) | 7 (12%) | 12,182 (13%) | 26,197 (34%) | 210 (33%) | 540 (15%) | 1,632 (49%) | 27 (47%) |
| Fever^2^ | 3,821 (2.9%) | 3,826 (4.8%) | 5 (8.5%) | 2,939 (3.1%) | 13,344 (17%) | 121 (19%) | 172 (4.8%) | 1,111 (33%) | 15 (26%) |
| Chills | 2,468 (1.9%) | 2,520 (3.2%) | 5 (8.5%) | 3,546 (3.8%) | 13,886 (18%) | 111 (17%) | 195 (5.5%) | 1,125 (34%) | 16 (28%) |
| Fatigue | 14,507 (11%) | 10,330 (13%) | 9 (15%) | 15,753 (17%) | 29,259 (37%) | 234 (36%) | 702 (20%) | 1,754 (53%) | 29 (51%) |
| Gastrointestinal^3^ | 3,865 (2.9%) | 3,002 (3.8%) | 2 (3.4%) | 3,319 (3.5%) | 8,472 (11%) | 78 (12%) | 180 (5.0%) | 703 (21%) | 13 (23%) |
| ^1^n (%); ^2^Fever defined as a temperature >38°C; ^3^Gastrointestinal symptoms include nausea, vomiting, diarrhoea and abdominal pain. | | | | | | | | | |
